# Evaluation of PainWaive: A consumer-grade EEG headset for remotely delivered neurofeedback and monitoring in chronic pain

**DOI:** 10.64898/2026.03.05.26347650

**Authors:** Nahian S Chowdhury, Joshua Rawsthorne, Negin Hesam-Shariati, Yann Quidé, Angus McIntyre, Sebastian Restrepo, Kevin Chen, Chin-Teng Lin, Toby Newton John, James W Middleton, Ashley Craig, Mark P Jensen, James McAuley, Sylvia M Gustin

## Abstract

Affordable home-based electroencephalography (EEG) headsets could widen access to EEG assessment, but require rigorous validation before research or clinical use. Here, we evaluated a custom-developed 2-channel sensorimotor headset (PainWaive) intended for remote neuro-feedback and longitudinal monitoring in chronic pain. Eighty participants (47 female; mean age 24.0 years, SD 7.9) completed two resting-state sessions with PainWaive and a research-grade 64-channel EEG system (LiveAmp), under eyes-open (EO) and eyes-closed (EC) conditions. Alpha, beta and theta power and peak alpha frequency (PAF) were derived from homologous sensorimotor channels (C1/C2). Relative reliability was quantified with intraclass correlation coefficients (ICCs), absolute reliability with SEM%, and cross-device consistency with between-device ICCs and Pearson correlations of overall spectral shape. ICCs/correlations were interpreted using pre-specified thresholds: fair 0.20–0.39, moderate 0.40–0.59, good 0.60–0.79, excellent ≥0.80. PainWaive and LiveAmp showed comparable absolute reliability across metrics (similar SEM%). Under EC, PainWaive reliability was excellent for alpha (0.81), theta (0.85) and PAF (0.94), and good for beta (0.72). Under EO, reliability was excellent for alpha (0.82), good for beta and PAF (0.61–0.72), and moderate for theta (0.59). Spectral-shape correlations between devices were excellent (r>0.90). Cross-device ICCs were good under EC for alpha/theta/PAF (ICC=0.66–0.77) though fair for beta (0.35). Under EO, ICCs were good for alpha (0.62), moderate for PAF (0.53), and fair for beta/theta (0.26–0.32). To assess performance under real-world use, we additionally analysed 2 clinical samples of individuals (total n = 8) with chronic pain who each completed 20 home-based neurofeedback sessions using PainWaive (160 sessions total). Within-session stability was good-to-excellent across metrics (ICCs>0.72). Overall, our findings suggest PainWaive is a reliable tool for the assessment of EEG metrics, supporting its use in research and clinical applications.

---

Electroencephalography (EEG) is widely used in research and clinical practice to study brain function and to diagnose conditions such as epilepsy and sleep disorders^1^. It is also used therapeutically in the form of neurofeedback (NFB), where individuals learn to modulate their brain activity in real time, providing a non-pharmacological option for conditions characterised by dysregulated oscillations, such as chronic pain^2,3^. However, traditional NFB applications have relied on research-grade EEG systems, which require lengthy setup, precise electrode placement and trained staff. This creates substantial barriers to wider clinical use^4–7^, particularly for people with impaired mobility, chronic illness, or those living in rural and remote regions^6–8^.

Portable, wireless, home-based EEG headsets offer a scalable alternative for NFB applications. However, most available consumer-grade systems target frontal or temporal regions for applications such as meditation, attention or sleep^9–17^, with relatively little focus on sensorimotor activity, which has been shown to be altered in chronic pain populations^18–20^. As such, there is currently no available consumer-grade sensorimotor EEG headset that has undergone rigorous test–retest and cross-device evaluation. Despite the challenges posed by condition-dependent variability in EEG spectral measures (e.g., eyes-closed vs eyes-open)^15–17,21–28^ and the greater inherent stability of some metrics compared to others (e.g. peak alpha frequency (PAF) is more stable than beta or theta power^29–33^), such evaluation is essential before widespread clinical and home-based implementation. To address this gap, we developed and evaluated PainWaive, an in-house 2-channel headset purpose-built to record sensorimotor activity from C1 and C2, designed for neurofeedback applications in people with chronic pain.

Since the PainWaive headset is intended for NFB, its evaluation must build on and improve the existing evidence base for testing consumer-grade EEG headsets. Indeed, several studies have examined test–retest reliability and cross-device consistency in consumer-grade EEG systems (mostly frontal/temporal channels). However, findings remain inconclusive due to the following methodological and technical limitations. Firstly, agreement with a research-grade system and test–retest reliability of the consumer device are often assessed in separate samples or under a single recording condition^13,15–17,23,25–28,34–39^. Cannard et al.^12^ reported strong correlations between the consumer-grade MUSE EEG headset and a research-grade system for alpha (r = 0.87) and theta power (r = 0.73) during eyes-closed resting-state recordings. In contrast, Mikhaylov et al^40^ found negligible correlations during eyes-open recordings (alpha r = –0.06, theta r = 0.12). It is unclear the extent to which these substantially lower correlations reflect the use of eyes-open conditions, device performance or an interaction between the two. Second, many studies lack statistical rigour, with few reporting *a priori* power or sample-size calculations^9,16,25,27,39,41,42^ and most summarising reliability only with relative indices (e.g., intraclass correlation coefficient, ICC), without absolute indices such as the standard error of measurement (SEM) or SEM%^9,16,25,27,39,41,42^. As such, it is unclear whether between-session variability is small enough to conclude clinically meaningful changes^43^. Finally, device-level technological limitations constrain achievable reliability: many low-cost headsets lack impedance monitoring^9–17^ and rely on dry electrodes, which reduce signal-to-noise ratios and increase susceptibility to motion artefacts compared with gel- or saline-based sensors^44–52^.

In designing the present evaluation of PainWaive, we therefore explicitly addressed these limitations by (a) assessing both test–retest reliability and cross-device consistency, (b) including both eyes-closed (EC) and eyes-open (EO) resting conditions in the same powered sample, and (c) incorporating impedance monitoring and saline-soaked electrodes to mitigate common technical constraints. Resting-state EEG under EC and EO conditions was recorded using both PainWaive and a research-grade system (Brain Products LiveAmp) in separate blocks across two sessions on the same day, with alpha, beta and theta power and PAF as primary outcomes. To evaluate intended real-world use, we also examined reliability in a sample of adults with chronic neuropathic pain who completed a 20-session PainWaive NFB program at home, using repeated recordings per session to assess the consistency of these metrics over time.

Based on expected differences in stability across metrics (PAF and alpha power typically more stable than beta and theta power)^53–61^ and recording conditions (EC typically more stable than EO), we hypothesised that, EC recordings would show excellent test–retest reliability (ICC ≥ 0.80) for alpha power and PAF, and good-to-excellent reliability (ICC = 0.60– 0.90) for beta and theta power. Under EO conditions, we predicted good-to-excellent reliability (ICC = 0.60–0.8) for alpha power and PAF and moderate-to-good reliability (ICC = 0.40– 0.7) for beta and theta power. We further predicted that cross-device consistency with the LiveAmp would be constrained by inherent reliability profiles. For EC, we expected alpha power and PAF to show good cross-device consistency (ICC = 0.60–0.8), with beta and theta power showing only moderate-to-good consistency (ICC = 0.40–0.70). For EO, we anticipated moderate-to-good cross-device consistency (ICC = 0.50–0.70) for alpha power and PAF, and fair-to-moderate consistency (ICC = 0.30–0.60) for beta and theta power.

## Methods

### Participants

#### Experimental Sample

Eighty participants (47 female, mean age = 24, SD = 7.9) were recruited from the University of New South Wales (UNSW) Sydney via two research participation platforms. Eligible participants were aged ≥18 years and able to provide informed consent; no additional exclusion criteria were applied. Psychology undergraduates were recruited through the SONA-1 system and received course credit. Additional participants, not limited to psychology students, were recruited through the SONA-P system and received monetary ($50) compensation. All provided written informed consent, and the study was approved by the UNSW Human Research Ethics Advisory Panel (iRECS#8344). Participation was voluntary, with the option to withdraw at any time.

Power analysis primarily focused on ICC, as it is the most conservative and widely reported index in EEG reliability research. The required sample size was calculated using G*Power 3.1 and was based on the principle of sample required to detect a target ICC metric against a null hypothesis minimum value^62^. We anticipated that the PainWaive system would show ICCs spanning a broad range, from fair-to-moderate (0.30–0.60) for eyes-open beta and theta cross-device consistency (representing the least consistent pairing), to good-to-excellent (0.8-0.99) for eyes-closed alpha power and PAF test–retest reliability (representing the most consistent pairing). For each sample size calculation, the null hypothesis was set at the lower bound of the relevant reliability category (e.g., ICC = 0.30 for beta/theta cross-device consistency), and the alternative hypothesis at the corresponding target value (e.g., ICC = 0.60 for beta/theta cross-device consistency). Using these assumptions, with a two-tailed α of 0.05 and 80% power, the largest sample size calculated was 56 participants (for detecting an ICC of 0.60 vs. a null of 0.30). To further increase power to 90% across all metrics and to safeguard against potential data loss, the final target sample size was set at 80 participants. Importantly, this study had no exclusion criteria due to the clinical safety of EEG and based on recommendations of the Ethics Advisory Panel.

#### Clinical Sample

Eight adults with chronic neuropathic pain were recruited as part of two separate single case experimental design studies: four with neuropathic eye pain and four with chronic low back pain. Each cohort completed a single-case experimental design (SCED) with multiple baselines, testing 20 sessions of home-based, self-directed EEG neurofeedback using the PainWaive headset. Participants acted as their own controls with daily clinical outcomes recorded across baseline, intervention, post-intervention, and follow-up phases. Participants and researchers were not blinded. This report focuses solely on the reliability of EEG metrics recorded during the intervention phase pooled across both cohorts; clinical outcomes for the eye pain cohort have been published previously,^63^ and clinical outcomes for the low back pain cohort will be reported separately.

Participants with neuropathic eye pain were 18–80 years old, had a clinical diagnosis of ocular neuropathic pain for ≥6 months, average pain ≥3/10 over the past week, were fluent in English, and had no planned changes to treatment or ocular procedures during the 12-week trial. Participants with chronic low back pain were ≥18 years, living in Australia, had ongoing low-back pain for ≥3 months, and average pain ≥4/10 over the past week. Exclusion criteria included severe neurodegenerative or psychiatric disorders, planned pain surgery, concurrent trial participation, or implanted cardiac/neurological devices. All procedures received institutional ethics approval, and all participants provided written informed consent. Studies were approved by the UNSW Human Research Ethics Advisory Panel (#iRECS8134 and HC230351)

### Protocol

#### Experimental Sample

A 2×2×2 repeated-measures design was used, with 2 headsets (PainWaive, LiveAmp), 2 conditions (EC, EO), and 2 sessions. Headset and task order were randomised for each participant using a computer-generated schedule, with counterbalancing across the sample to ensure equal representation of all sequences.

#### Clinical Sample

Each of the four participants from each cohort were randomly allocated (computer-generated) to staggered baseline lengths of 7, 10, 14, or 17 days, with lengths differing by ≥3 days. All participants from each cohort began baseline on the same day. The intervention comprised 20 home-based EEG neurofeedback sessions over 4 weeks.

### Data Collection Procedures – Experimental Sample

#### Pre-Session Instructions

Participants were instructed to avoid hair products and caffeine before their session to optimise electrode conductivity and recording quality.

#### Preparation of LiveAmp

Prior to recording, the headset and electrode sponges were soaked in a saline solution. EEG was recorded with BrainVision Recorder (v1.27.0001, Brain Products, Germany) at a 250 Hz sampling rate using the 64-channel Live-Amp headset (Fig. 1a). The LiveAmp was arranged according to the international 10-20 system (Fig. 1b), with the ground electrode at FPz and the reference at FCz. Electrodes C1 and C2, from which key spectral measures were derived, were carefully monitored to meet this threshold, and ensure high signal quality, matching criteria between LiveAmp and PainWaive. Impedance was kept below 50 kΩ, a threshold supported by previous research and the LiveAmp manual as unlikely to cause signal loss^64,65^.

**Figure 1:**
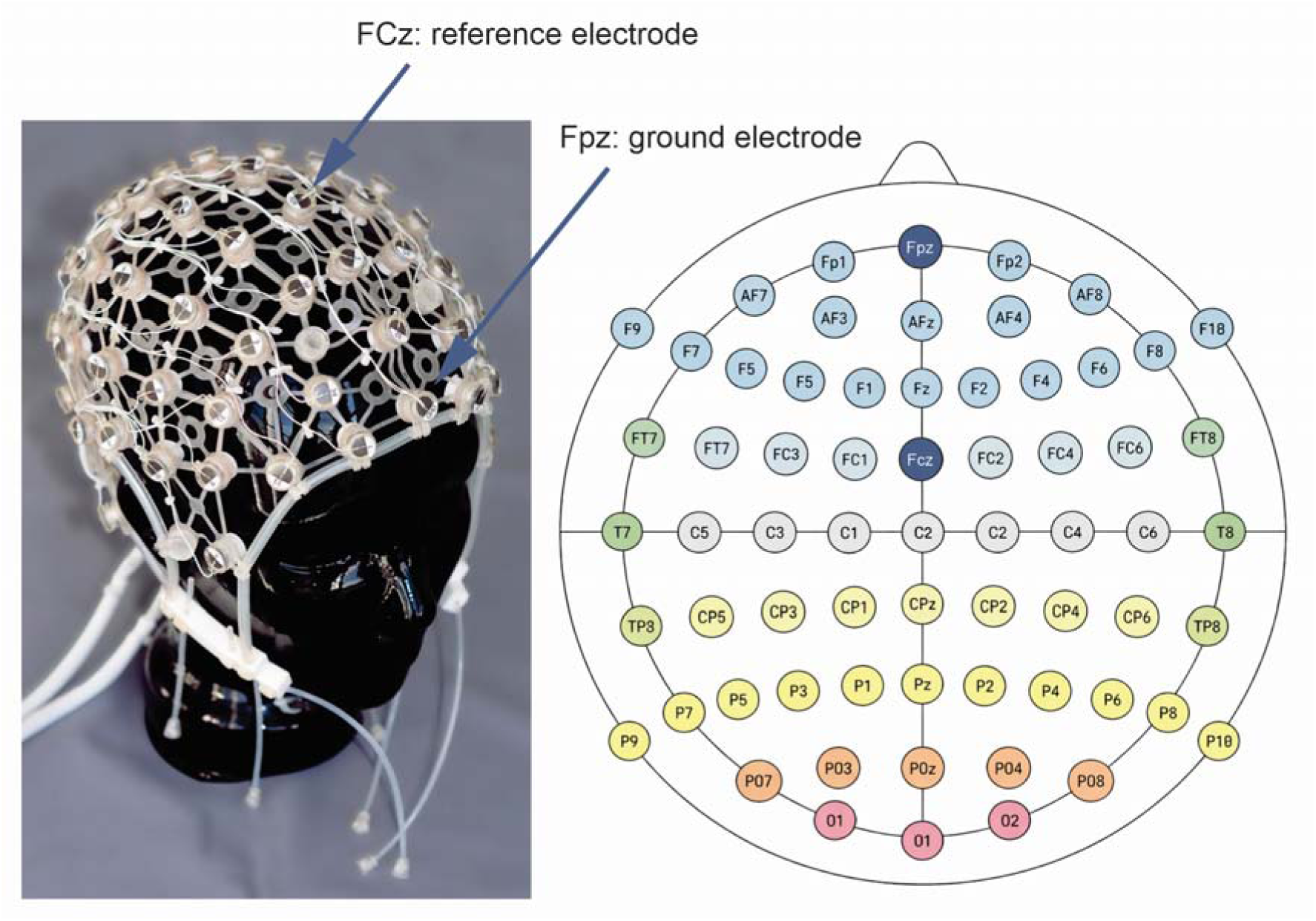
Live-Amp headset and electrode configuration. **a)** LiveAmp EEG headset used for data collection. **b)** 64-channel electrode layout based on the international 10-20 system, which positions electrodes at standardised intervals across the scalp to ensure consistent coverage of cortical regions. Reference electrodes (FCz, red arrow) and ground electrodes (FPz, green arrow) are indicated in both panels. Images sourced from Brain Products, Germany (https://www.brainproducts.com/<u>), with modifications made using Microsoft PowerPoint.</u>

#### Preparation of the PainWaive

Before recording, electrode sponges and ear clips were soaked in a saline solution. EEG was collected using the PainWaive headset (Fig. 2) and OpenBCI GUI (v5.2.0, OpenBCI, USA) at a sampling rate of 200 Hz. The headset had two recording silver chloride electrodes mounted in a 3D-printed frame positioned at C1 and C2, in accordance with the international 10-20 system, with ground and reference electrodes integrated into left and right earclips^2^. Small Hydro-Link sponges are used to complete the connection between the scalp and the electrodes. Details about the PainWaive headset are provided in the supplementary materials. Impedances were maintained below 50 kΩ, to match the LiveAmp set-up using the OpenBCI GUI (v5.2.0).

**Figure 2:**
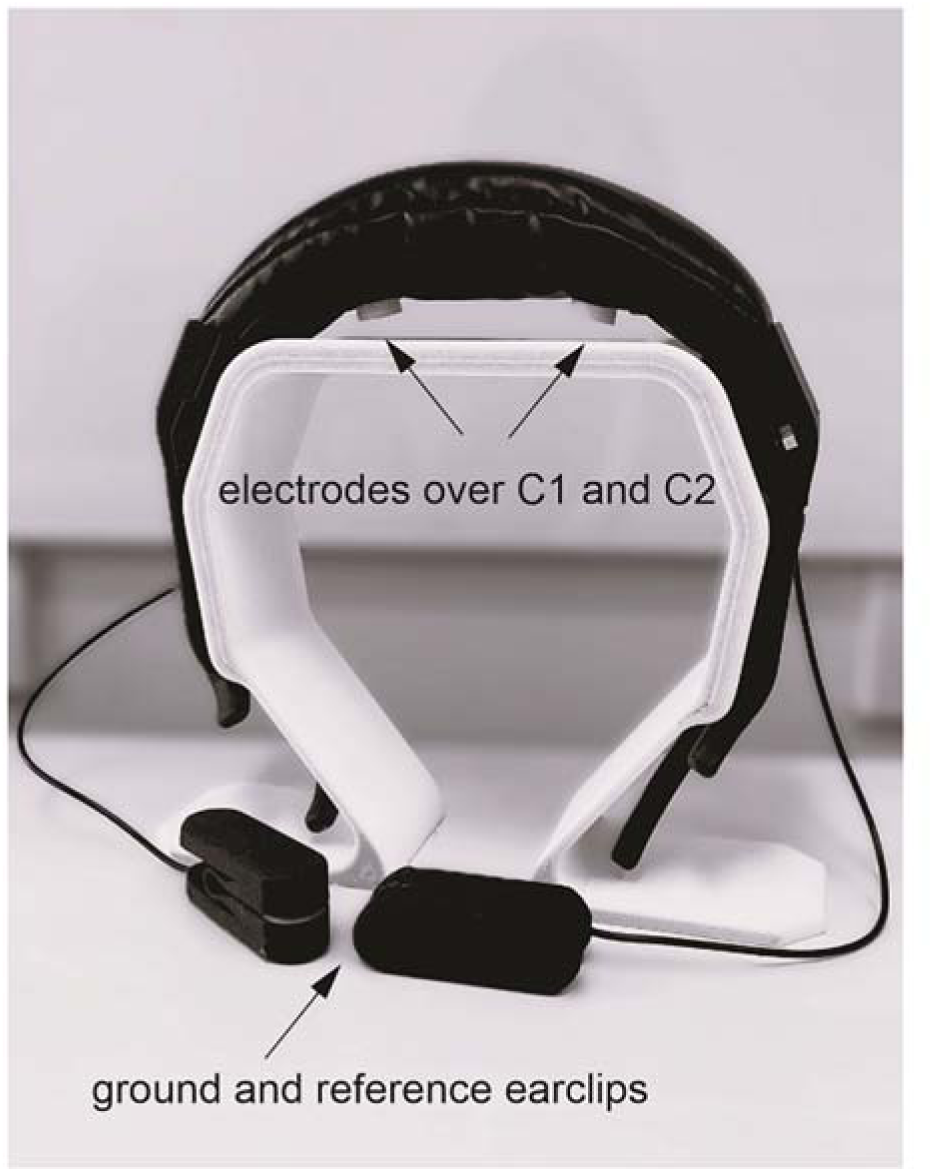
PainWaive headset. Arrows indicate recording electrodes over C1 and C2, and reference and ground earclips

#### Calibration and Recording Procedure

Before each recording session, participants completed a standardised sequence of movements and facial actions (eyes up/down, left/right, blinking, jaw clenching, and 30 seconds of eye closure) to identify artefact and noisy channels. The recording protocol included two 3-minute tasks: (a) fixation on a central cross with eyes open (EO condition) and (b) eyes closed while remaining awake (EC condition). Each task was completed twice with both headsets, with session and device order randomised and counterbalanced across participants.

### Data Collection Procedures – Clinical Sample

During each home session, EEG was recorded from C1 and C2 over the sensorimotor cortex after verifying impedances ^63^. Sessions began with a 2-minute EO resting state EEG; the relative power in theta, alpha, and beta bands set the initial game thresholds. Participants then completed five x 2.5 minutes neurofeedback rounds designed to suppress theta (4–7 Hz) and high-beta (20–30 Hz) while reinforcing high alpha/low beta (10–15 Hz). Four types of game scenarios provided visual feedback via background colour and character motion (Jellyfish, Plane, Bird and Rocket) with successful modulation making the flying object move and background brighten. Participants were instructed to minimise movement and use recommended strategies - positive thinking, focusing on game features, and slow breathing exercises to regulate the targeted bands; raw EEG was not displayed. Optional background music supported relaxation. Adherence and quality control were managed via a secure dashboard. At the start of each session, the tablet captured a headset-placement photo; EEG and de-identified photos were automatically uploaded and linked to participant IDs. The research team reviewed uploads after each session to confirm placement and screen for artefacts, contacting participants with troubleshooting as needed^63^

### EEG Data Pre-Processing Pipeline

Data were pre-processed using MATLAB (vR2024b), EEGLAB (v2023_1), and FieldTrip (v20200215). LiveAmp recordings were down-sampled from 250 Hz to 200 Hz to match the PainWaive. Device comparisons focused solely on the C1 and C2 channels to match the PainWaive montage. All data were re-referenced to the C1-C2 average to harmonise reference schemes and avoid bias from differing reference electrode locations, consistent with previous studies comparing home-based headsets to research-grade systems^9,12,14,36,37,40,66–69^. Noisy channels were rejected if their variance exceeded the channel mean by >3 standard deviations; however, C1 and C2 were always retained because care was taken to allow direct comparisons between devices without the need for interpolation or imputation. Data were band-pass filtered (2-45 Hz) and segmented into 5-second epochs, with epochs exceeding ±150 µV removed.

### EEG Data Post-Processing Pipeline

Raw EEG data were analysed offline using a Fast Fourier Transform (FFT) with a Hanning window to reduce edge artefacts and decompose the signal into frequency components. Power spectral density (PSD) was computed at 0.2 Hz intervals with 0% epoch overlap. Spectral power from C1 and C2 was averaged across channels and trials to obtain a mean frequency profile per participant. Power values were z-transformed within participants to normalise spectral profiles for between-subject comparison while preserving within-participant spectral shape. Band powers for theta (4-7 Hz), alpha (8-12 Hz), and beta (13-30 Hz) were calculated by summing the z-transformed power within each frequency bin. To further characterise individual alpha activity, PAF was calculated using the Centre of Gravity formula.

### Statistical Analysis

#### Descriptives (Experimental Sample)

We computed means and SDs per session, condition and headset. Normality was assessed using skewness and kurtosis. Distributions with skewness between ±2 and kurtosis below 3.29 were considered approximately normal for medium-sized samples, consistent with guidelines^70,71^. All analyses were conducted in JASP (v0.95.1).

#### Absolute Test-Retest Reliability (Experimental Sample)

Absolute reliability, which reflects measurement error and within-individual variability, was assessed using the standard error of measurement (SEMeas); lower SEMeas values indicate better measurement precision and reduced within-individual variability^72,73^. The following formula was used: SEMeas=√mean squared error, where the mean squared error (or ‘residual error’) was obtained from one-way repeated measures analysis of variance applied to test and retest measurements^74^. SEMeas%, was also calculated (using the formula: SEMeas%= SEMeas/pooled mean*100, where pooled mean was obtained from both testing sessions), to indicate the relative size of the measurement error^75,76^

#### Relative Test-retest reliability (Experimental Sample)

Relative test–retest reliability was quantified using intraclass correlation coefficients (ICC) based on a two-way mixed-effects, single-measurement, consistency model, following the Shrout & Fleiss and McGraw & Wong conventions^77,78^. This model assumes that both subjects and measurement occasions are fixed and that each participant is measured under the same conditions. The ICC quantifies the proportion of the total variance that is attributable to true between-subject differences relative to total variance. ICCs were computed in MATLAB (R2024b) using a custom script and interpreted according to established thresholds^79,80^, including poor (≤0.2), fair (0.2-0.39), moderate (0.4-0.59), good (0.60-0.79) and excellent (0.8-0.99).

#### Relative Test-Retest Reliability in other channels (Experimental Sample)

To determine whether poor reliability at C1/C2 might simply reflect a particularly noisy scalp region, we also calculated test–retest reliability for all 64 channels of the LiveAmp system. For each channel and EEG metric, we estimated two-way mixed-effects, single-measure, consistency ICCs, generating topographical maps of relative reliability across the scalp.

#### Cross-Device Consistency (Experimental Sample)

Cross-device consistency between the 2-channel device and the 64-channel LiveAmp was quantified using ICCs based on a two-way random-effects, single-measurement, absolute-agreement model, again following Shrout & Fleiss and McGraw & Wong^77,78^. This model assumes that both devices are sampled from a larger population of possible measurement systems and assesses absolute agreement between them. To further characterise cross-device agreement, Pearson correlation coefficients (r) were calculated between the full spectral profiles recorded with each headset to assess the similarity of the spectral shape. Together, these indices provide a comprehensive assessment of both individual rankings (ICC) and the similarity of spectral distributions (r) across devices.

#### Relative Test-Retest Reliability (Clinical Sample)

Participants across both SCEDs completed 20 home-based neurofeedback sessions (total 160 sessions), each comprising five recordings (‘games’), with each game separated by ∼1 minute. We treated each neurofeedback session as a separate sampling point and asked: *Do the five recordings within a session yield similar EEG values (alpha, beta, theta power and PAF)?* To answer this, we pooled all available sessions across participants and constructed a session × recordings matrix for each metric (rows = sessions, columns = Games 1–5). We then computed a two-way mixed-effects, single-measurement, consistency ICC. In this framework, sessions are the targets and games are repeated measurements. The resulting pooled ICC for each metric therefore reflects the extent to which a single recording provides a stable estimate of the session’s EEG value under the same conditions, and is interpreted as our primary device-performance index.

## Results

### Descriptives (Experimental Sample)

Tables 1 and 2 present descriptive statistics for each headset, session and condition. Most measures met normality criteria based on skewness and kurtosis. Exceptions included theta power in both LiveAmp sessions and beta and theta power in the PainWaive, which exceeded kurtosis thresholds. In the EO condition (Table 2), several LiveAmp measures, notably alpha power and PAF, also showed elevated skewness and kurtosis, with PainWaive alpha power likewise deviating from normality. As the majority of metrics satisfied normality assumptions, data were not transformed and non-parametric tests were not applied, in order to preserve interpretability and allow direct comparisons across devices and conditions.

**Table 1:** Descriptive statistics for the LiveAmp and PainWaive spectral measures during eyes-closed (EC) testing. The table reports mean, standard deviation (SD), skewness, and kurtosis for alpha power, beta power, theta power, and PAF across two sessions (S1, S2). Distributions were considered normal when skewness values fell between ±2 and kurtosis values were below 3.29. Values that did not meet the assumption of normality are marked with an asterisk (*)

| **LiveAmp** |  |  |  |  |  |  |  |  |
| --- | --- | --- | --- | --- | --- | --- | --- | --- |
|  | Alpha Power<br>S1 | Beta Power<br>S1 | Theta<br>Power S1 | PAF S1 | Alpha<br>Power<br>S2 | Beta<br>Power<br>S2 | Theta<br>Power S2 | PAF S2 |
| Mean | 12.33 | -24.69 | 10.89 | 9.960 | 12.77 | -25.15 | 11.90 | 9.971 |
| SD | 14.68 | 3.297 | 6.425 | 0.264 | 14.83 | 3.028 | 7.042 | 0.284 |
| Skewness | 1.531 | -0.027 | 1.882 | -0.723 | 1.430 | -0.568 | 1.915 | -1.063 |
| Kurtosis | 1.213 | 1.364 | 4.193* | 1.333 | 0.914 | 0.046 | 5.075* | 1.802 |
| **PainWaive** |  |  |  |  |  |  |  |  |
|  | Alpha Power<br>S1 | Beta Power<br>S1 | Theta<br>Power S1 | PAF S1 | Alpha<br>Power<br>S2 | Beta<br>Power<br>S2 | Theta<br>Power S2 | PAF S2 |
| Mean | 17.10 | -28.31 | 13.70 | 10.05 | 15.50 | -28.91 | 14.11 | 10.04 |
| SD | 13.31 | 4.355 | 7.081 | 0.325 | 12.63 | 5.002 | 7.261 | 0.327 |
| Skewness | 0.835 | 1.327 | 1.948 | -1.244 | 0.838 | 0.974 | 1.534 | -1.144 |
| Kurtosis | -0.466 | 5.312* | 5.00* | 0.525 | -0.164 | 6.575* | 3.901* | 0.576 |

**Table 2:**
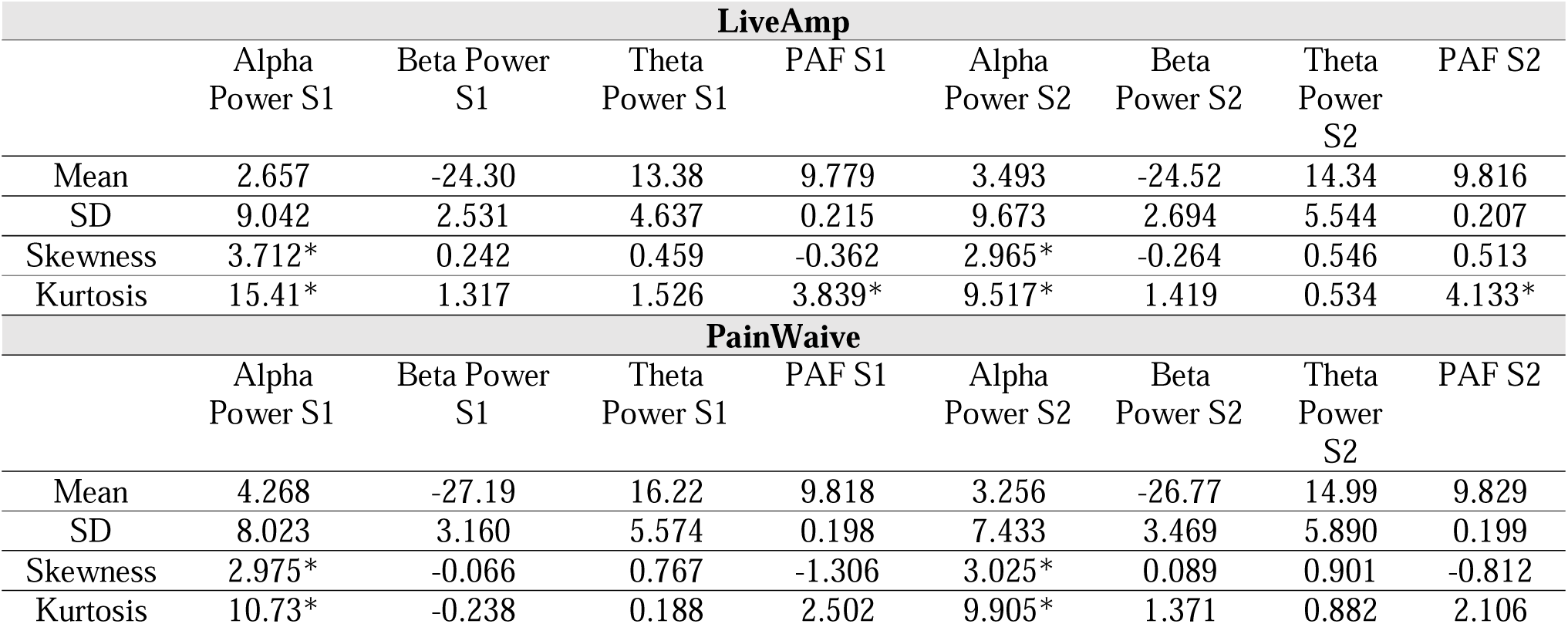
Descriptive statistics and kurtosis for LiveAmp and PainWaive spectral measures during eyes-open (EO) testing. The table reports mean, standard deviation (SD), skewness, and kurtosis for alpha power, beta power, theta power, and PAF across two sessions (S1, S2). Distributions were considered normal when skewness values fell between ±2 and kurtosis values were below 3.29. Values that did not meet the assumption of normality are marked with an asterisk (*)

| **LiveAmp** |  |  |  |  |  |  |  |  |
| --- | --- | --- | --- | --- | --- | --- | --- | --- |
|  | Alpha Power S1 | Beta Power S1 | Theta Power S1 | PAF S1 | Alpha Power S2 | Beta Power S2 | Theta Power S2 | PAF S2 |
| Mean | 2.657 | -24.30 | 13.38 | 9.779 | 3.493 | -24.52 | 14.34 | 9.816 |
| SD | 9.042 | 2.531 | 4.637 | 0.215 | 9.673 | 2.694 | 5.544 | 0.207 |
| Skewness | 3.712* | 0.242 | 0.459 | -0.362 | 2.965* | -0.264 | 0.546 | 0.513 |
| Kurtosis | 15.41* | 1.317 | 1.526 | 3.839* | 9.517* | 1.419 | 0.534 | 4.133* |
| **PainWaive** |  |  |  |  |  |  |  |  |
|  | Alpha Power S1 | Beta Power S1 | Theta Power S1 | PAF S1 | Alpha Power S2 | Beta Power S2 | Theta Power S2 | PAF S2 |
| Mean | 4.268 | -27.19 | 16.22 | 9.818 | 3.256 | -26.77 | 14.99 | 9.829 |
| SD | 8.023 | 3.160 | 5.574 | 0.198 | 7.433 | 3.469 | 5.890 | 0.199 |
| Skewness | 2.975* | -0.066 | 0.767 | -1.306 | 3.025* | 0.089 | 0.901 | -0.812 |
| Kurtosis | 10.73* | -0.238 | 0.188 | 2.502 | 9.905* | 1.371 | 0.882 | 2.106 |

### Absolute Reliability (Experimental Sample)

Table 3 reports SEMeas percentages, indexing between-session fluctuations for each metric. SEM% values were broadly similar across headsets, indicating that overall between-session variability was comparable for the LiveAmp and PainWaive.

**Table 3:** Absolute reliability of spectral measures expressed as standard error of the mean (as a percentage)

|  | **Eyes Closed** |  |  |  | **Eyes Open** |  |  |  |
| --- | --- | --- | --- | --- | --- | --- | --- | --- |
|  | Alpha Power | Beta Power | Theta Power | PAF | Alpha Power | Beta Power | Theta Power | PAF |
| **LiveAmp** | 15.5 | 4.7 | 13.95 | 0.37 | 35.4 | 3.6 | 11.86 | 0.61 |
| **PainWaive** | 15.4 | 4.5 | 11.2 | 0.46 | 39.8 | 4.2 | 13.3 | 0.60 |

### Relative test–retest reliability (Experimental Sample)

During EC testing (Fig. 3), relative reliability was excellent for alpha power, theta power and PAF across both devices, whereas beta power showed moderate reliability for the LiveAmp and good reliability for the PainWaive.

**Figure 3.**
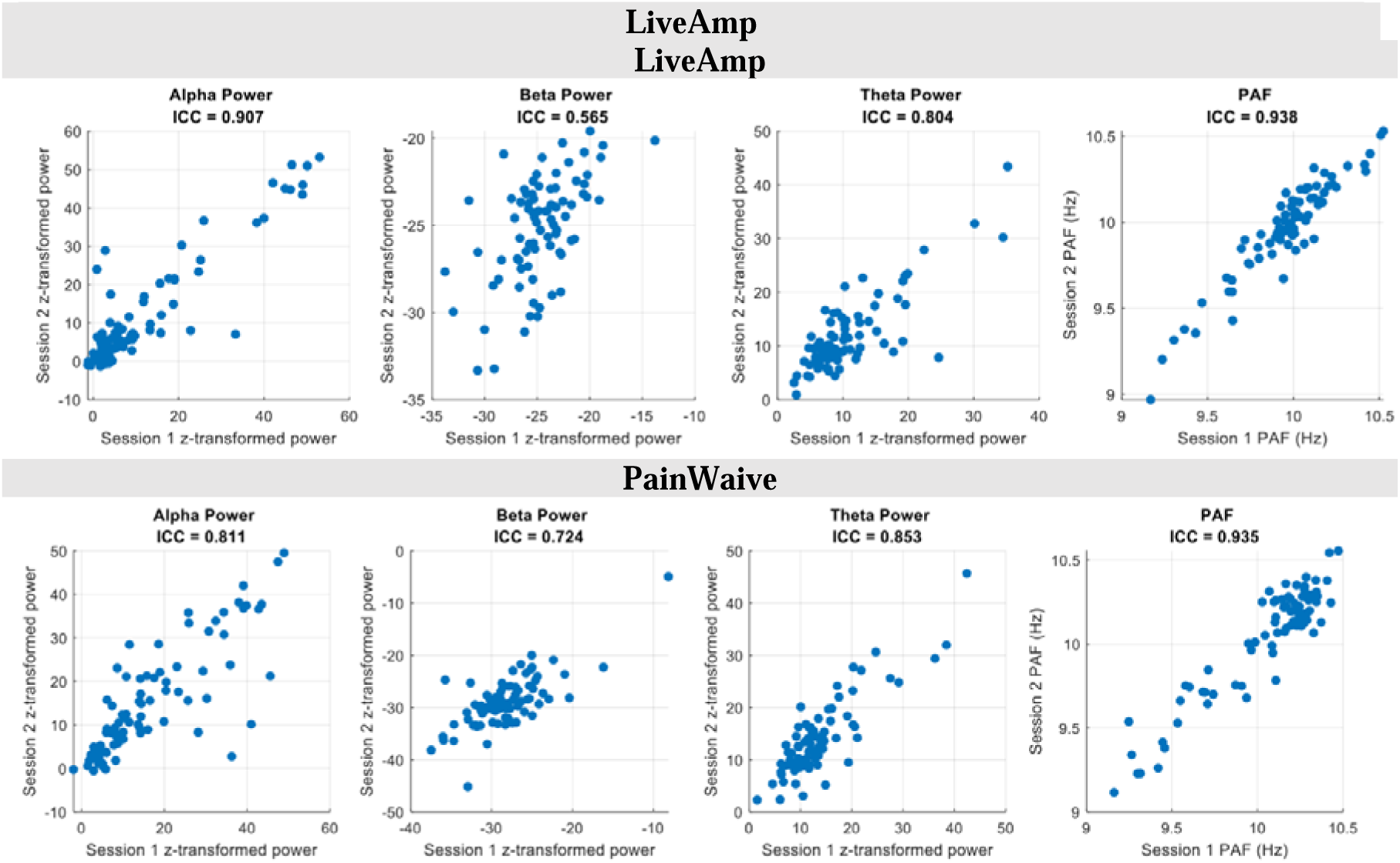
Relative test-retest reliability of spectral measures (ICC) during eyes-closed (EC) testing with the LiveAmp and PainWaive.

During EO testing (Fig. 4), the test-retest reliability of alpha power remained in the excellent range across devices, good range for beta and PAF. Reliability of theta power was in the moderate range for the PainWaive and good range for the LiveAmp.

**Figure 4:**
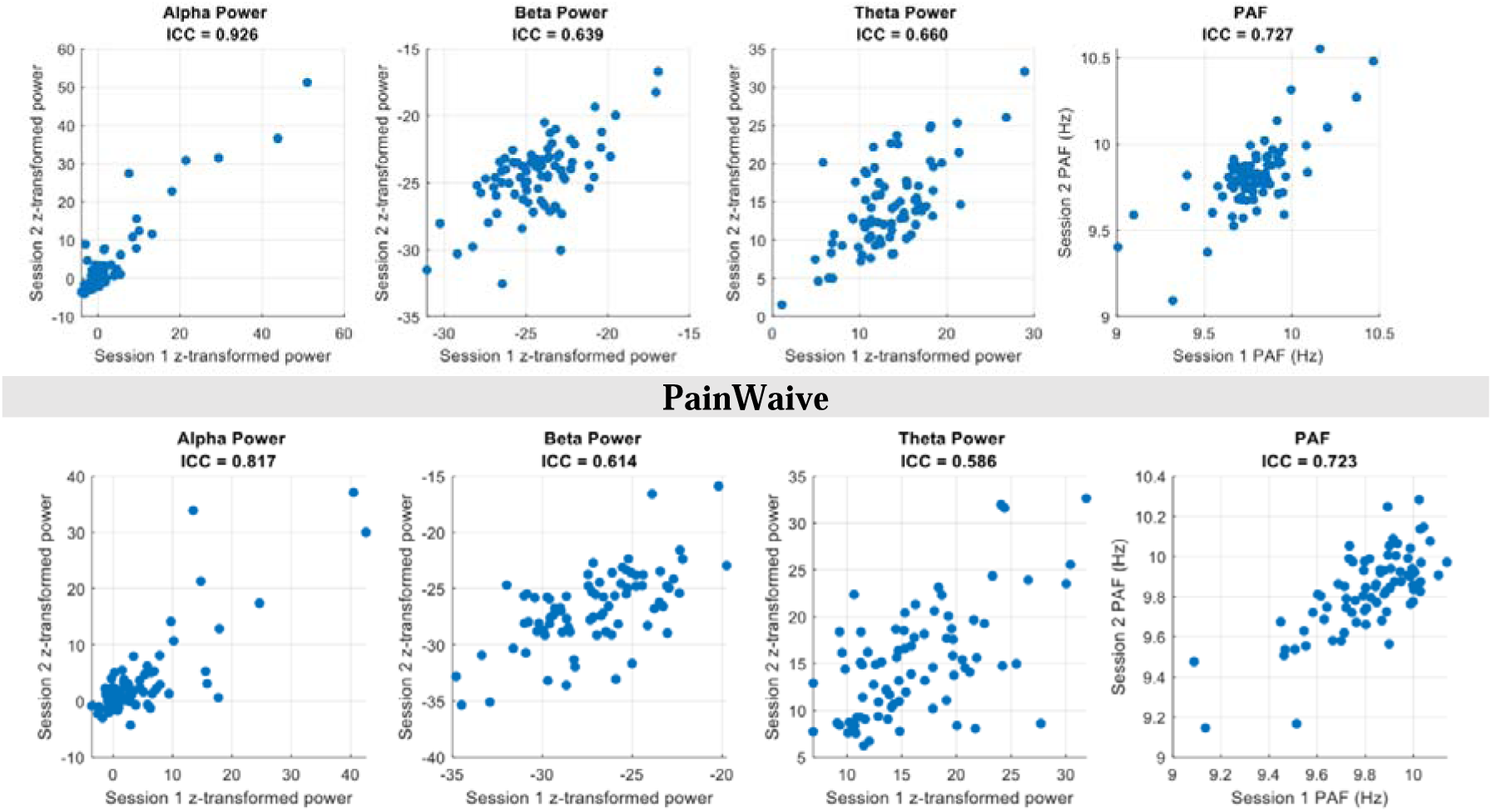
Relative test-retest reliability of spectral measures (ICC) during eyes-open (EO) testing with the LiveAmp and PainWaive.

### Relative Test-Retest Reliability in Other Channels (Experimental Sample)

Under EC conditions (Fig. 5, top), alpha power and PAF exhibited uniformly high ICC values across the scalp, with most channels in the good-to-excellent range. Theta power also showed broadly good reliability, with only small clusters of slightly lower ICCs over lateral regions. In contrast, beta power displayed a more heterogeneous pattern, with good reliability over posterior sites but only fair-to-moderate reliability over midline frontal–central channels.

**Figure 5:**
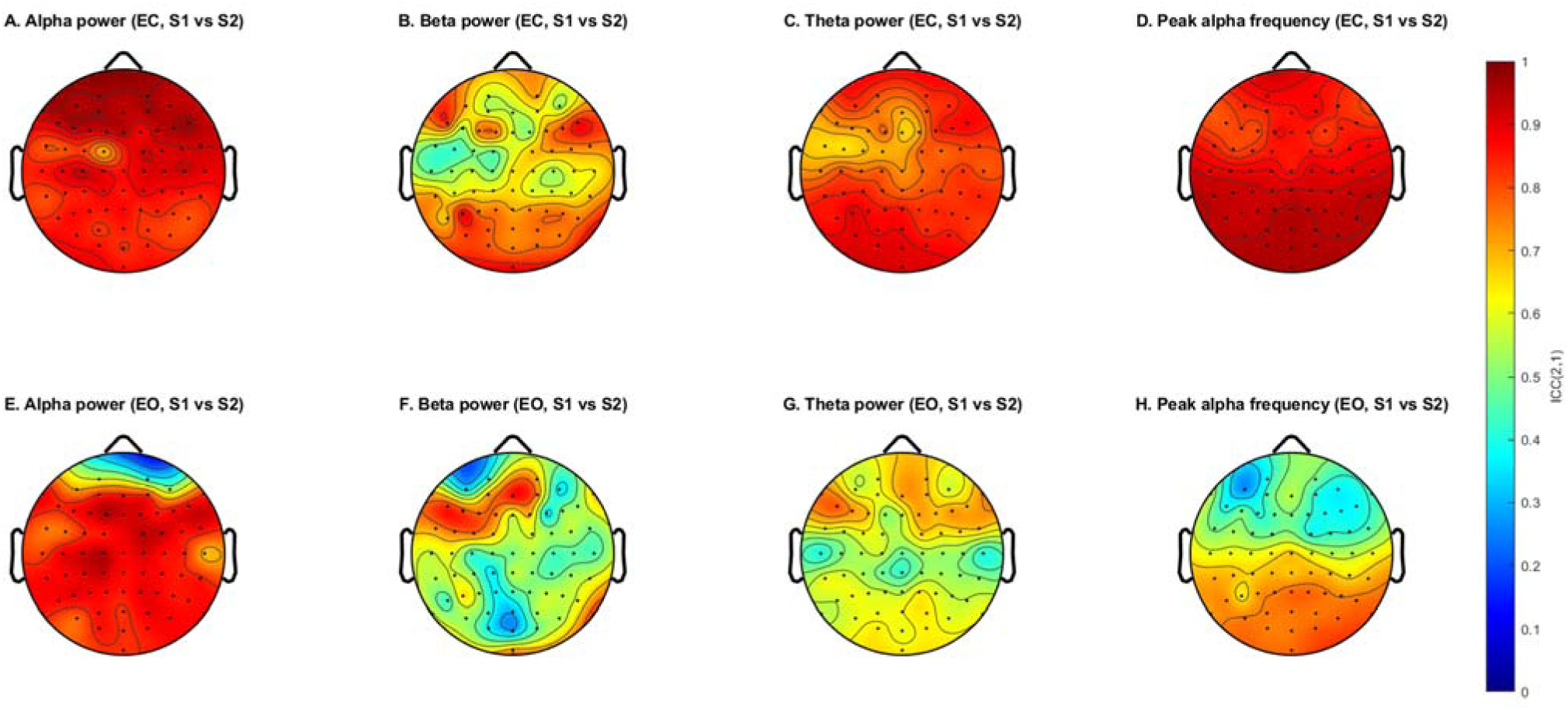
Relative test-retest reliability of EEG metrics from all channels recorded using the LiveAmp.

Under EO conditions (Fig. 5, bottom), alpha power remained highly reliable over central and posterior electrodes but showed reduced reliability at frontal sites. Theta and beta power showed the lowest and most spatially variable ICCs, with predominantly fair-to-moderate values over central and posterior regions and only isolated areas of good reliability. For PAF, EO reliability was still good-to-excellent over central and posterior channels but dropped sharply at frontal electrodes.

### Cross-Device Consistency (Experimental Sample)

Power spectral density plots during EC testing (Fig. 6, top) show highly similar spectral profiles for the PainWaive and LiveAmp across sessions, with excellent consistency in spectral values across frequency bins (r > .90) between devices. During EO testing (Fig. 6, bottom), spectral consistency remained excellent (r > .90). For band-limited metrics (Fig. 7), consistency during EC was in the good range for alpha power, theta power and PAF, and fair range for beta power. In EO, consistency was in the good range for alpha power, moderate range for PAF, and fair range for theta and beta power.

**Figure 6:**
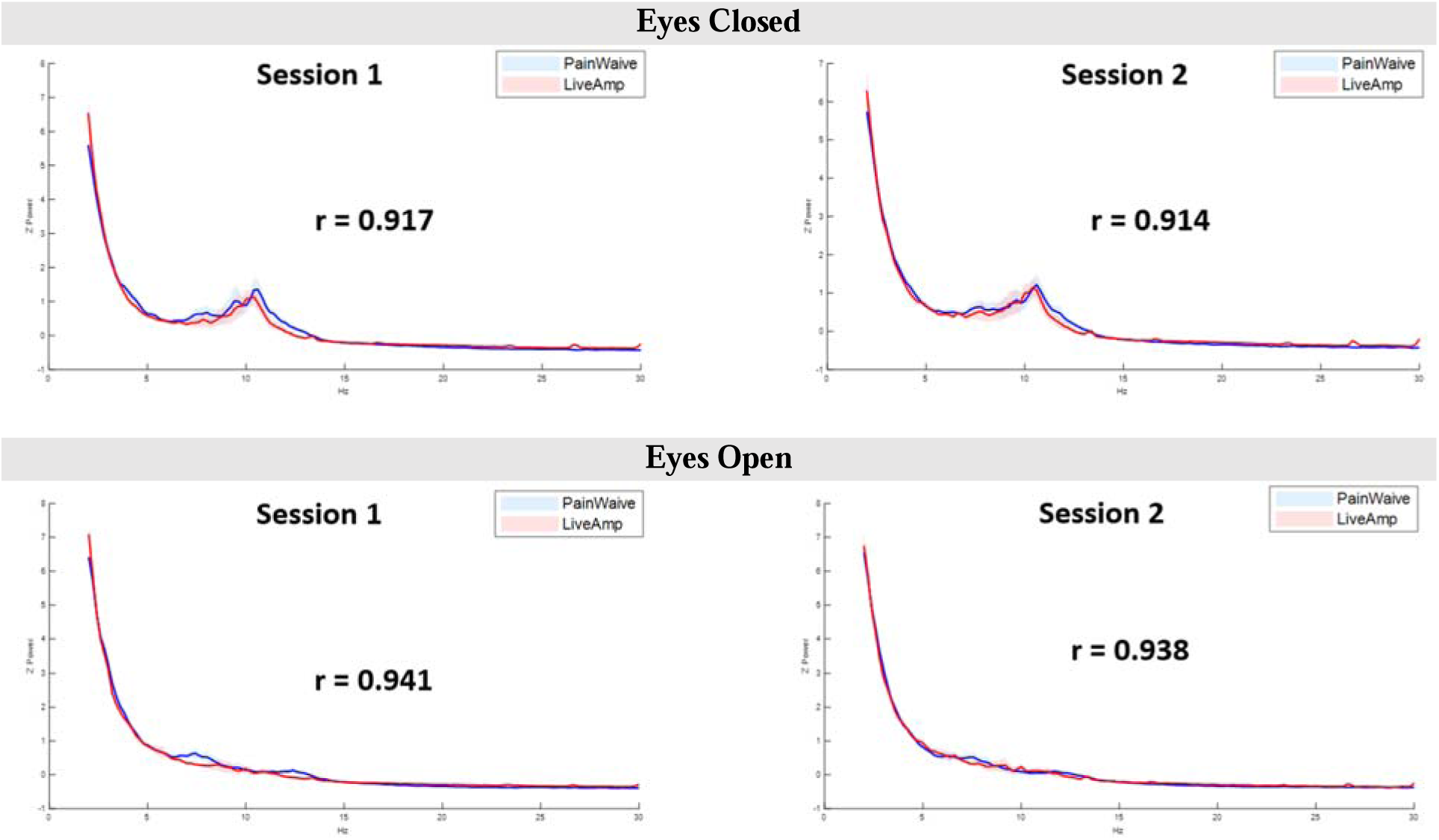
EEG spectral profile (r) across devices and conditions. Mean z-transformed power is shown for PainWaive (blue) and LiveAmp (red) with 95% CI shading.

**Figure 7:**
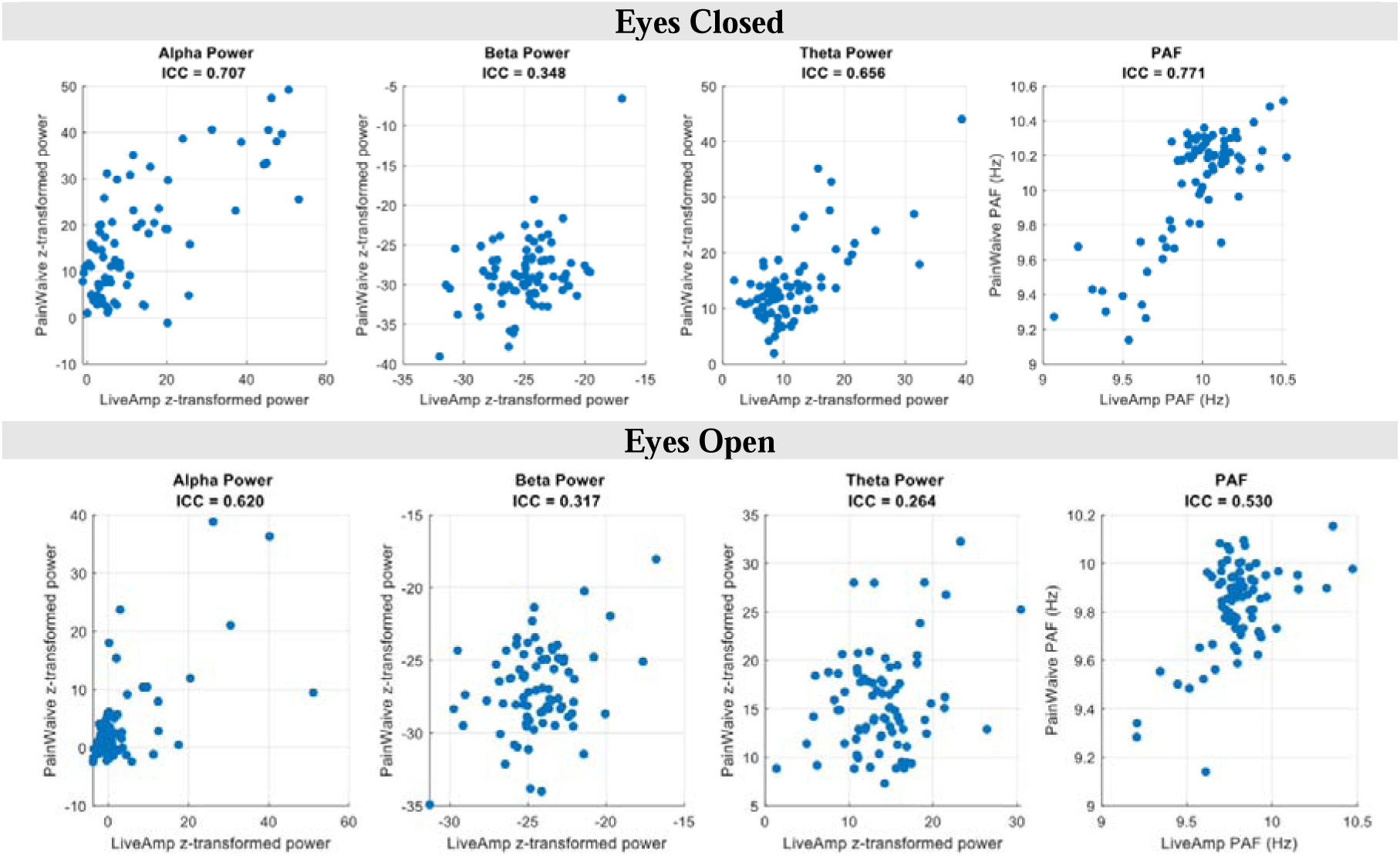
Relative validity of spectral measures (ICC) comparing the LiveAmp and PainWaive headsets.

### Relative Test-Retest Reliability (Clinical Sample)

In the clinical neurofeedback sample (Fig. 8), across all sessions completed by the eight participants, the consistency of alpha power, PAF, beta power and theta power across the five neurofeedback games within each session was in the good-to-excellent range (ICC > 0.72), indicating that a single recording provided a stable estimate of the session-level EEG value under nominally identical conditions

**Figure 8:**
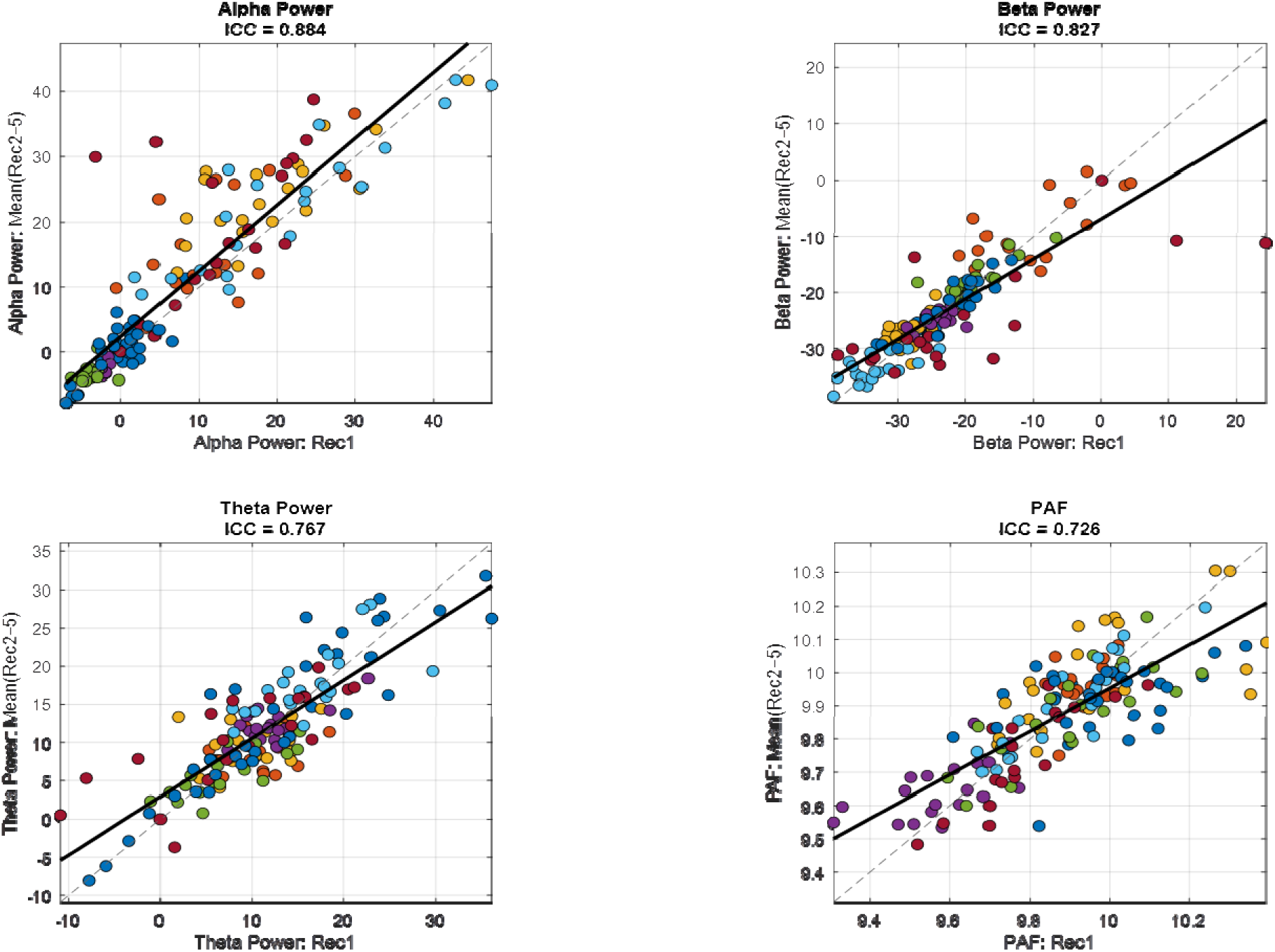
Relative test retest reliability of spectral measures (ICC) of the PainWaive in the Clinical sample. Each colour represents a different participant who completed 20 neurofeedback sessions, with x-axis showing the metric from recording 1, and y-axis showing the mean of the remaining 4 recording. The ICC on the top represents the agreement in values across the 5 games

### Supplementary Analysis

In the supplementary materials (Tables S1-S8), we present all findings as a function of different analytic pipelines, examining: (a) the use of log-transformed absolute power instead of z-scores, (b) DPSS versus Hanning tapers, and (c) a more relaxed cleaning approach involving minimal filtering versus the present rigorous pipeline. Across these analyses, the overall pattern of results remained similar for alpha power and PAF, both of which consistently showed good-to-excellent relative test-retest reliability and moderate-to-good cross-device consistency. However, beta and theta results varied more as a function of analytic choices. Pipelines using log-transformed spectral values produced the strongest beta and theta cross-device estimates (particularly for EC beta and EO theta), whereas pipelines using z-scores (chosen in the primary analysis) showed weaker validity, especially in eyes-open recordings. Substituting Hanning tapers with DPSS tapers tended to enhance reliability for several metrics, but this did not translate to improved beta or theta cross-device consistency, which remained low for eyes-open data in z-transformed DPSS pipelines.

## Discussion

This study aimed to evaluate the test–retest reliability and cross-device consistency of PainWaive, an in-house EEG headset recording from sensorimotor electrodes, developed for neurofeedback applications in chronic pain. PainWaive is intended to enable remote neuro-feedback delivery and longitudinal digital biomarker monitoring outside the laboratory, which requires evidence of measurement stability, interoperability with research-grade systems, and robustness to analytic variation. To address key limitations of prior consumer EEG research, we (a) evaluated test–retest reliability and cross-device consistency under both eyes-closed (EC) and eyes-open (EO) conditions, (b) extended analyses to additional channels and a clinical sample, (c) reported absolute reliability metrics and a priori sample size calculations, and (d) developed a headset with impedance monitoring and saline-soaked electrodes rather than dry sensors. We found that, across metrics, relative reliability ranged from good to excellent for EC recordings and from moderate to excellent for EO recordings, with absolute reliability comparable between devices. Test–retest reliability was also good to excellent in the clinical sample of participants completing neurofeedback sessions, supporting the stability of PainWaive-derived metrics in both research and applied settings. Cross-device consistency for spectral shape was excellent under both EC and EO. For band-limited metrics, EC recordings showed good agreement for alpha power, PAF and theta power, and fair agreement for beta power. Under EO, consistency was moderate-to-good for alpha power and PAF, and fair for theta and beta power. Overall, our study provides a comprehensive evaluation of a home-based EEG headset designed for chronic pain neurofeedback.

### Test-retest reliability

The achievable reliability of EEG metrics is constrained not only by device performance but also inherent fluctuations of different metrics (with alpha power and PAF typically showing greater stability) and from recording condition (EC typically greater stability than EO). Acknowledging this, we found good reliability for beta power and excellent reliability for PAF, alpha, and theta under EC conditions. Under EO conditions, reliability was excellent for alpha, good for PAF and beta, and moderate for theta. These values fall within the typical reliability range reported for these metrics and conditions^53–61^. The observed values were also comparable to those obtained with the research-grade LiveAmp system. Thus, the PainWaive headset shows an expected level of reliability for an EEG device. As most consumer-grade headsets have focused on frontal recordings, this study also illustrates the performance of an in-house EEG device developed for sensorimotor measurements. Our findings suggest potential reliability advantages of using impedance monitoring and wet electrodes. However, because we did not directly compare PainWaive with a dry-electrode system, definitive conclusions regarding the performance of devices with and without impedance monitoring and wet electrodes cannot be drawn.

Our assessment of absolute reliability using SEM% provides a useful contribution to the home-based EEG literature, as many prior studies do not report this metric^9,16,25,27,39,41,42^. Here we show that the between-session absolute fluctuations across metrics were comparable with the PainWaive and the LiveAmp, further supporting the similarity in performance between the headsets. Beyond reliability benchmarking, SEM% estimates clarify the minimal detectable difference for each spectral measure, which is essential for intervention design and sample size calculations. Collectively, these results provide valuable information for future studies using home-based EEG systems.

A key limitation of using a laboratory sample to assess device performance is that testing is conducted under optimal conditions that include: trained experimenters, controlled environments, and immediate quality feedback. Real-world home use is likely to introduce additional variability, including less precise electrode placement, reduced impedance control, environmental noise, and variable adherence. To bridge this gap, we also examined test– retest reliability of the same metrics across neurofeedback sessions conducted in two separate clinical studies, therefore reflecting the intended real-world application of the device. Within a given session, all metrics showed good-to-excellent consistency, further strengthening our conclusions regarding the reliability of the PainWaive headset.

### Cross-device consistency

Assessment of cross-device consistency is inherently constrained by the reliability of the underlying spectral metrics themselves, differences in headset setup (e.g., montage, reference, channel count, and the influence of electrical signals from other channels), and by the variability introduced through reliability assessments. It is also important to note that, while research-grade systems are treated as a gold standard, these systems are still subject to impedance variation, environmental noise, and other artefacts^30,33^. This suggests that observed estimates of cross-device consistency do not necessarily reflect device performance alone but also broader physiological and methodological variability.

Acknowledging these constraints, PainWaive showed excellent agreement in overall spectral profile (*r*’s > 0.9), with closely matched amplitude–frequency patterns across devices. This suggests that the overall shape of the EEG spectrum is highly reproducible between PainWaive and the research-grade system. In other words, PainWaive does not appear to selectively boost or attenuate specific frequencies (e.g., over-damping high beta or exaggerating alpha). Preservation of spectral shape across devices suggests that these metrics are less likely to reflect device-specific artefacts and more likely to index underlying neurophysiology. This, in turn, indicates that PainWaive and the research-grade system may be used interchangeably for biomarkers that depend primarily on overall spectral shape ^81,82^.

For band-limited power metrics, cross-device consistency followed the same band- and condition-dependent pattern observed for reliability. Under EC, ICCs were good for alpha, theta, and PAF and fair for beta, whereas under EO the ICCs were good for alpha, moderate for PAF, and fair for beta and theta. Although lower cross-device consistency is expected during EO recordings as shown by Mikhaylov et al.^40^ who reported poor-to-fair agreement between MUSE 1 and ActiChamp Plus (alpha: r = –0.06; beta: r = 0.07; theta: r = 0.12), other work has found higher beta-band consistency during EC recordings than observed in our study. For example, Cannard et al.^12^ reported excellent cross-device agreement in EC beta power between consumer and research-grade systems, with correlations of r = 0.84. One possible explanation for these differences is a reference mismatch between headsets: LiveAmp used FCz as the online reference, whereas PainWaive used an earlobe reference. Cannard et al.^12^ similarly showed that reference choice matters: when a wearable system was compared against a research-grade system using a different reference (average reference), agreement dropped for some measures, consistent with reference-dependent differences in spectral estimates. Another factor may be sensor location: Cannard et al. tested a MUSE headset for frontal channel, whereas in our LiveAmp data, reliability across channels was generally lower over the sensorimotor region, which is recorded by PainWaive. A final consideration relates to signal scaling. In the present study, our primary analyses used z-transformed (i.e., relative) power, which normalises each participant’s spectral profile. This may compress between-band differences and reduce the apparent contribution of higher-frequency activity (e.g., beta) relative to alpha. By contrast, log-transformed absolute power avoids this relative scaling and produced noticeably stronger beta and theta agreement in our supplementary pipelines, suggesting that transformation choice may contribute to variability in cross-device estimates. Overall, further research is required to clarify the influence of pipeline, montage and sensor location in the reliability and cross-device consistency estimates for consumer-grade headsets.

### Broader implications

This work robustly evaluated home-based EEG headsets across multiple experimental conditions within a single study. As mentioned, many studies do not simultaneously assess absolute and relative reliability and cross-device consistency for both eyes-closed and eyes-open recordings^13,15–17,23,25–28,34–39^, with Lau-Zhu et al. highlighting the lack of systematic evaluations of home-based EEG devices as a major barrier to translation^83^. As such, the current study provides a reproducible evaluation framework and reference values which may improve the comparability across studies.

The clinical implications of our findings are especially encouraging for neurofeedback applications targeting band-limited activity over sensorimotor cortex. PainWaive showed excellent agreement in overall spectral shape, good-to-excellent test–retest reliability in experimental and clinical samples, and moderate-to-good cross-device validity for alpha metrics.

While theta and beta showed weaker cross-device agreement (especially eyes-open), they remained reliable within PainWaive. Together, these results support PainWaive as a feasible platform for longitudinal, home-based neurofeedback and remote monitoring in chronic pain, where within-device stability and reliable targets are most critical.

### Limitations and future directions

Two limitations require acknowledgement. First, there is no true ‘ground truth’ EEG signal in our study. In particular, the combination of preserved within-device reliability and imperfect cross-device agreement for some metrics (especially under EO) makes it difficult to determine which device, if either, is closer to the underlying physiological ‘bullseye’. Future work should adopt multimodal validation approaches, for example, combining structural MRI–based forward models^84^ with scalp EEG to test how well different devices recover cortical sources, or comparing scalp and intracranial EEG as a stronger physiological benchmark.

Second, the technological differences between PainWaive and LiveAmp are non-trivial. LiveAmp provides higher channel density and more sophisticated hardware, whereas PainWaive prioritises accessibility and ease of use. This asymmetry inherently favours the research-grade system in direct device-to-device comparisons. Moving forward, it may be more informative to ask whether home-based devices are ‘good enough’ for their intended functional outcomes (e.g., pain reduction, adherence, engagement) rather than focusing solely on how closely they match lab-grade systems on spectral metrics.

### Conclusion

In summary, PainWaive demonstrated moderate-to-excellent test–retest reliability across EC and EO conditions, excellent cross-device consistency in overall spectral shape, and good-to-moderate cross-device consistency for PAF and alpha power. For beta and theta bands, cross-device agreement was fair to moderate in the primary pipeline, although supplementary analyses showed that moderate validity can be achieved under some analytic configurations, particularly for eyes-closed data. These findings suggest that straightforward one-to-one interchangeability with research-grade systems such as LiveAmp should not be assumed without carefully matched montages, electrode layouts, and processing choices. Overall, the results support the use of PainWaive for research and clinical applications.

## Acknowledgements

The authors would like to thank Lorimer Moseley for his helpful comments on early versions of the manuscript.

## Data Availability

De-identified data are available to qualified researchers on reasonable request, in accordance with the ethics approvals and study protocols governing this research.

## Author Contributions

N.S.C: Conceptualization, data curation, formal analysis, investigation, methodology, software, validation, visualization, writing (original draft), and writing (reviewing and editing).

J.R: Conceptualization, data curation, formal analysis, investigation, methodology, software, validation, visualization, writing (original draft), and writing (reviewing and editing).

N.H.S: Conceptualization, validation, visualization, and writing (reviewing and editing).

Y.Q. Conceptualization, validation, visualization, and writing (reviewing and editing).

A.M: Software/Hardware, and writing (reviewing and editing).

S.R: Software/Hardware, and writing (reviewing and editing).

K.C: Software/Hardware, and writing (reviewing and editing).

C.T.L: Funding acquisition, and writing (reviewing and editing).

T.N.J: Funding acquisition, and writing (reviewing and editing).

J.M. Funding acquisition, and writing (reviewing and editing).

A.C. Funding acquisition, and writing (reviewing and editing).

M.J. Funding acquisition, and writing (reviewing and editing).

J.A. Funding acquisition, and writing (reviewing and editing).

S.M.G. Funding acquisition, supervision, validation, visualization, writing (original draft), and writing (reviewing and editing).

