## Supplementary Material for "Evaluation of PainWaive: A consumer-grade EEG headset for remotely delivered neurofeedback and monitoring in chronic pain"

**Supplementary Material for Evaluation of PainWaive: A Consumer-Grade Headset for assessing sensorimotor electroencephalographic activity**

**Supplementary Methods**

We assessed test–retest reliability (within-device ICC) and cross-device consistency (LiveAmp vs PainWaive ICC) under multiple plausible analysis pipelines, varying (i) taper (Hanning vs DPSS), (ii) cleaning stringency (relaxed vs standard), and (iii) transformation of absolute power (log vs z). These choices were selected because each represents a defensible alternative in resting-state spectral analysis that can affect estimated power, signal-to-noise ratio, and between-session stability.

### Spectral estimation: single-taper Hanning vs multi-taper DPSS

**Hanning (single-taper fast Fourier transform).** In the primary pipeline, spectral power was estimated using a conventional FFT applied to 5-s epochs after multiplying each epoch by a Hanning window. The Hanning taper gradually reduces the signal toward zero at epoch edges, which reduces edge discontinuities and therefore limits **spectral leakage** (i.e., power from a true frequency “spilling” into neighbouring bins). This approach is widely used, computationally simple, and provides relatively high frequency resolution for a given epoch length, but can exhibit higher variance in the PSD estimate because it relies on a single tapered estimate per epoch.

**DPSS (multitaper FFT).** As an alternative, we recomputed PSD using a multitaper approach based on **discrete prolate spheroidal sequences (DPSS; Slepian tapers).** DPSS tapers are designed to maximise energy concentration within a defined bandwidth, providing controlled spectral smoothing while minimising leakage outside that bandwidth. In practice, multitapering generates multiple independent tapered PSD estimates per epoch and averages them, which typically **reduces estimator variance** and improves robustness to noise and transient artefacts, at the cost of some spectral smoothing (i.e., slightly reduced frequency specificity).

### Cleaning stringency: standard vs relaxed pre-processing

**Standard cleaning.** This matched the main manuscript pipeline: data were band-pass filtered (2–45 Hz), segmented into 5-s epochs, and artefactual epochs were rejected if amplitudes exceeded ±150 µV. Channels were flagged as noisy if variance exceeded the channel mean by >3 SD; however, **C1 and C2 were always retained** to preserve direct cross-device comparability without interpolation or imputation. This approach prioritises removal of high-amplitude, non-physiological activity likely to inflate broadband power and reduce between-session stability.

**Relaxed cleaning.** In the relaxed pipeline, we retained the same segmentation and spectral estimation steps but **did not perform epoch rejection** and **did not perform noisy-channel rejection** (beyond the enforced retention of C1 and C2, which applied across pipelines). This analysis tests the “minimal handling” scenario that may better reflect pragmatic home recordings or large-scale datasets where aggressive rejection is undesirable. Because artefact contamination can systematically bias absolute power and increase PSD variance, we expected relaxed cleaning to particularly affect higher frequencies and absolute-power metrics.

### Transformations of power: log-scaled absolute power vs z-normalised spectra

**Log transformation (absolute power).** For log-power pipelines, power spectra density (PSD) was retained on an absolute scale (e.g., µV²/Hz) and transformed using a logarithmic transform (log10). Log-scaling compresses the heavy-tailed distribution of power values, reduces heteroscedasticity. Band power was then derived by averaging or summing the log-PSD within canonical frequency ranges (theta 4–7 Hz; alpha 8–12 Hz; beta 13–30 Hz).

**Z transformation (within-participant spectral normalisation).** For z-power pipelines, PSD values were z-transformed **within participant** across frequency bins to normalise overall magnitude while preserving each participant’s **relative spectral profile** (spectral shape). This emphasises features such as relative alpha prominence over broadband amplitude differences (e.g., due to impedance, coupling, or amplifier gain). After z-transformation, band power was computed by summing z-values within the same theta/alpha/beta frequency bins. This approach is particularly relevant for cross-device comparisons where absolute scaling may differ, but device-agnostic spectral shape may be more stable.

**Supplementary Results**

Supplementary Tables S1 to S8 show the test-retest reliability and cross-device consistency results of the different pipelines

PAF (especially eyes-closed) showed excellent within-device reliability for both systems across all pipelines (typically ICC ≈ 0.93–0.95) and consistently good cross-device consistency (eyes-closed cross-device consistency ≈ 0.77 across pipelines).

Alpha power showed good-to-excellent reliability in both systems across pipelines, and moderate-to-good cross-device consistency (typically ~0.55–0.75 depending on state and pipeline).

Beta and theta cross-device consistency were the most pipeline-dependent, with effects most pronounced in eyes-open recordings. Under some pipelines, beta/theta cross-device consistency was moderate, whereas under others it dropped into the fair range. In contrast, within-device reliability for beta/theta was more stable than cross-device consistency, however still showed some dependence on transform/taper (e.g., certain z-transform/Hanning configurations reduced beta reliability, while DPSS-based pipelines tended to stabilise reliability).

Overall, the sensitivity analyses suggest that PainWaive’s reliability and cross-device consistency are less variant to pipeline for alpha-band and PAF, with conclusions largely unchanged across reasonable analytic choices. However, beta and theta agreement between devices was more variant depending on analytic decisions, particularly for eyes-open data, indicating that these bands may be more susceptible to differences in sensor noise characteristics, spectral leakage/estimation choices, and normalisation/transformation Cleaning stringency (relaxed vs standard) generally produced small changes relative to the effects of taper/transform; i.e., the overall pattern of results did not depend strongly on whether cleaning was relaxed or standard.

**Table S1. Z-transformed · Hanning Taper · Standard cleaning (Primary pipeline).** LA = LiveAmp, PW = PainWaive.

| **Metric** | **EC Reliability (LA / PW)** | **EC Cross-device consistency** | **EO Reliability (LA / PW)** | **EO Cross-device consistency** |
| --- | --- | --- | --- | --- |
| Alpha | 0.922 / 0.817 | 0.748 | 0.915 / 0.795 | 0.581 |
| Beta | 0.776 / 0.675 | 0.511 | 0.793 / 0.681 | 0.320 |
| Theta | 0.845 / 0.882 | 0.687 | 0.752 / 0.669 | 0.312 |
| PAF | 0.944 / 0.945 | 0.776 | 0.780 / 0.781 | 0.521 |

**Table S2. Z-transformed · Hanning Taper · Minimal cleaning**

| **Metric** | **EC Reliability (LA / PW)** | **EC Cross-device consistency** | **EO Reliability (LA / PW)** | **EO Cross-device consistency** |
| --- | --- | --- | --- | --- |
| Alpha | 0.903 / 0.812 | 0.702 | 0.925 / 0.821 | 0.601 |
| Beta | 0.594 / 0.700 | 0.359 | 0.599 / 0.599 | 0.310 |
| Theta | 0.827 / 0.846 | 0.644 | 0.652 / 0.578 | 0.256 |
| PAF | 0.933 / 0.934 | 0.770 | 0.694 / 0.726 | 0.524 |

**Table S3. Z-transformed · DPSS · Minimal cleaning**

| **Metric** | **EC Reliability (LA / PW)** | **EC Cross-device consistency** | **EO Reliability (LA / PW)** | **EO Cross-device consistency** |
| --- | --- | --- | --- | --- |
| Alpha | 0.912 / 0.817 | 0.758 | 0.939 / 0.801 | 0.553 |
| Beta | 0.837 / 0.687 | 0.577 | 0.834 / 0.683 | 0.319 |
| Theta | 0.874 / 0.885 | 0.704 | 0.771 / 0.695 | 0.328 |
| PAF | 0.937 / 0.945 | 0.773 | 0.756 / 0.779 | 0.503 |

**Table S4. Z-transformed · DPSS · Standard cleaning**

| **Metric** | **EC Reliability (LA / PW)** | **EC Cross-device consistency** | **EO Reliability (LA / PW)** | **EO Cross-device consistency** |
| --- | --- | --- | --- | --- |
| Alpha | 0.921 / 0.817 | 0.748 | 0.915 / 0.795 | 0.581 |
| Beta | 0.837 / 0.687 | 0.577 | 0.834 / 0.683 | 0.319 |
| Theta | 0.874 / 0.885 | 0.704 | 0.771 / 0.695 | 0.328 |
| PAF | 0.937 / 0.945 | 0.773 | 0.756 / 0.779 | 0.503 |

**Table S5. Log-transformed · Hanning · Minimal cleaning**

| Metric | EC Reliability (LA / PW) | EC Cross-device consistency | EO Reliability (LA / PW) | EO Cross-device consistency |
| --- | --- | --- | --- | --- |
| Alpha | 0.878 / 0.803 | 0.597 | 0.871 / 0.718 | 0.554 |
| Beta | 0.784 / 0.859 | 0.639 | 0.738 / 0.799 | 0.540 |
| Theta | 0.736 / 0.700 | 0.383 | 0.756 / 0.469 | 0.555 |
| PAF | 0.933 / 0.934 | 0.770 | 0.694 / 0.726 | 0.524 |

**Table S6. Log-transformed · Hanning · Standard cleaning**

| Metric | EC Reliability (LA / PW) | EC Cross-device consistency | EO Reliability (LA / PW) | EO Cross-device consistency |
| --- | --- | --- | --- | --- |
| Alpha | 0.882 / 0.811 | 0.597 | 0.890 / 0.717 | 0.576 |
| Beta | 0.794 / 0.863 | 0.639 | 0.721 / 0.799 | 0.540 |
| Theta | 0.733 / 0.708 | 0.393 | 0.745 / 0.477 | 0.525 |
| PAF | 0.938 / 0.935 | 0.771 | 0.728 / 0.723 | 0.530 |

**Table S7. Log-transformed · DPSS · Minimal cleaning**

| Metric | EC Reliability (LA / PW) | EC Cross-device consistency | EO Reliability (LA / PW) | EO Cross-device consistency |
| --- | --- | --- | --- | --- |
| Alpha | 0.884 / 0.809 | 0.613 | 0.896 / 0.685 | 0.561 |
| Beta | 0.821 / 0.805 | 0.587 | 0.809 / 0.761 | 0.530 |
| Theta | 0.755 / 0.762 | 0.426 | 0.817 / 0.413 | 0.479 |
| PAF | 0.937 / 0.945 | 0.773 | 0.756 / 0.779 | 0.503 |

**Table S8. Log-transformed · DPSS · Standard cleaning**

| Metric | EC Reliability (LA / PW) | EC Cross-device consistency | EO Reliability (LA / PW) | EO Cross-device consistency |
| --- | --- | --- | --- | --- |
| Alpha | 0.884 / 0.813 | 0.611 | 0.896 / 0.672 | 0.579 |
| Beta | 0.833 / 0.808 | 0.594 | 0.796 / 0.758 | 0.537 |
| Theta | 0.772 / 0.765 | 0.450 | 0.820 / 0.429 | 0.503 |
| PAF | 0.944 / 0.945 | 0.776 | 0.780 / 0.781 | 0.521 |
